# Wearable Focal Muscle Vibration Improves Upper Limb Function in People with Sub-acute Stroke

**DOI:** 10.1101/2024.11.11.24317091

**Authors:** Imran Khan Niazi, Imran Amjad, Irum Farooq, Hina Shafi, Usman Rashid, Nitika Kumari, Nusratnaaz Shaikh, Mads Jochumsen, Kelly Holt, Heidi Haavik, Simon F. Farmer, Amit N. Pujari

## Abstract

The objective of the study was to compare the effects of two focal muscle vibration (FMV) stimulation frequencies (60 Hz and 120 Hz), combined with conventional physical therapy (PT), on upper limb impairment and function in people with sub-acute stroke when FMV is delivered through a wearable FMV device.

The study was a parallel group, randomized controlled trial conducted at the Rehabilitation Centre of Railway General Hospital, Rawalpindi, Pakistan. A total of 98 individuals with sub-acute stroke were randomly allocated to control group (eight weeks of conventional physical therapy, (PT)) or experimental groups (eight weeks of conventional physical therapy combined with focal muscle vibration (FMV) at 60 Hz (PT + FMV60 Hz) or 120 Hz (PT + FMV120 Hz)). Outcome measures included Fugl Meyer Scale for upper extremity (FMUE), Motor Assessment Scale (MAS), and Modified Ashworth Scale (MASh). They were collected at baseline and after eight weeks of treatment. Data were analysed using linear regression model. The study was registered on the National Institutes of Health ClinicalTrials.gov clinical trial registry (Identifier NCT04289766).

At post-intervention time-point, FMUE scores varied across groups (F (2, 81) = 7.2, p = 0.001), MAS scores did not vary across groups (F (2, 81) = 0.2, p = 0.8) and MASh rank changes varied across groups (F (2, 81) = 3.3, p = 0.04). There were no differences between the PT + FMV60 Hz and the PT + FMV120 Hz groups.

This study provides the evidence that wearable Focal Muscle Vibration (at 60 and 120 Hz) improves the motor outcome of sub-acute stroke patients. Thus, it can be used along with conventional physiotherapy as a valid intervention to promote upper limb function and reduce spasticity in sub-acute post-stroke patients.

## 1 Introduction

Stroke is one of the leading causes of disabilities in adults ^1–3^. Over 85% stroke survivors experience upper limb paresis ^4^ and around 50% stroke survivors suffer with upper limb discoordination with significant many unable to regain any functional use of their paretic upper limbs ^5,6^. Although some therapeutic approaches are available to help restore normal motor function in stroke survivors with upper limb impairments ^7–9^, quality of evidence and hence effectiveness of these interventions in improving upper limb motor function is still debated ^10,11^. Further, many of the current therapeutic approaches used in improving motor function post-stroke, focus on compensation and accommodation ^12–15^, instead of restoration to normal or near normal movement function. Partly due to the overall clinical consensus that patients showing severe impairment in the upper limbs 1-month post-stroke may not regain any normal function in the long-term ^14,16–18^.

It is well known that post-stroke, sensory systems are disrupted and this disruption has been thought to play major role in sensorimotor dysfunction of the paretic upper limb ^19^, including its impact on the ability to control functional arm movements^20^. As loss of proprioceptive and tactile sensation is prevalent consequence^19^ of stroke and affects control of the arm, regaining proprioceptive and tactile sensation, by targeting neuroplastic mechanisms, is one way to improve sensorimotor ability of the paretic limb. Indeed, studies show that repeated sensory stimulation positively affects motor cortical excitability ^21,22^. In this context, studies have shown the importance of somatosensory afferents in connecting with motor cortical circuits to bring about neuroplastic changes in the M1. With studies showing long term changes in functional improvement following the use of FMV (90 minutes over 3 days) in stroke population ^22^.

More recently, mechanical vibrations have been used as somatosensory stimulation and to improve motor function ^23^ and to treat post-stroke upper limb muscle spasticity ^24^. When vibration stimulus is used for exercise or physical rehabilitation, it can be broadly categorized into two types (a) stimulation directly applied to a specific muscle or tendon, and (b) indirect vibration which is not muscle-specific and is delivered either through the feet while standing on a platform or through the hands by holding a device. Direct vibration applied through muscle or tendon is also known as focal muscle vibration (FMV) or segmental vibration (SV) and sometimes also referred as repetitive Muscle Vibration (rMV) (5). Whereas, indirect vibration delivered through the hands is commonly referred to as upper limb vibration (ULV) ^25,26^ and indirect vibration delivered to the lower limbs is referred to as whole-body vibration (WBV) ^27,28^. The human body’s response to vibration stimuli is highly complex, with affects reported on neuromuscular, musculoskeletal and endocrine systems^29^. In the present study only neuromuscular mechanisms will be discussed. Readers are referred to extensive review for a more detailed treatment on other areas ^29^. From the neuromuscular perspective, direct vibration (i.e. focal or segmental vibration) of muscle or tendon alters the transmission of primary and secondary muscle afferents (Ia, Ib, and type II afferents) ^30–32^, along with modulation of cutaneous mechanoreceptors ^33^, and cortical excitability ^34^. Vibration stimulation’s ability to selectively excite and alter proprioceptive afferents and its connections to the CNS is of importance, as post-stroke changes can be observed in proprioception and tactile sensation and such changes are implicated in disrupted planning and control of limb posture and movement^35^. Further, along with the modulation in afferent transmission, when applied to the muscle-tendon complex, direct vibration produces an involuntary contraction in the vibrated muscle through the tonic vibration reflex ^32^ and vibration acts to inhibit the antagonist muscle activity, a useful mechanism to reduce hyperexcited stretch reflex observed in spasticity ^36^. However, the effects of vibration, including modulation of afferent transmission and Tonic Vibration Reflex (TVR) generation, are highly dependent upon the characteristics of the vibration stimulus being applied ^30–32,37,38^. These characteristics primarily include vibration location, frequency, amplitude, duration, muscle contraction and fatigue level and the protocol of vibration application ^28,39,40^. Vibration effects also depend upon individuals’ characteristics e.g.. age, gender and disability type ^41^. Thus, a range of factors need to be carefully considered when planning an effective vibration stimulation treatment regime with the aim of improving motor functions of stroke populations ^42^.

In recent years, FMV has been investigated as a modality to treat spasticity. Spasticity is defined as the velocity-dependent increase in muscle tone/resistance that may follow neurological disease or injury (e.g. stroke and spinal cord injury) ^43,44^. Although the precise underlying mechanisms leading to spasticity are still debated and not fully understood, the consensus is spasticity is largely the result of hyperexcitability of the stretch reflex ^43,44^. Therefore, a neuromodulatory signal which can reduce hyperexcitability of the stretch reflex may act to reduce spasticity ^45,46^. Both, direct and indirect vibration, have been shown to produce a marked reduction in the H-reflex drive, a reliable indicator that vibration stimuli reduces the hyperexcitability of the stretch reflex ^47,48^.

Further, direct vibration (i.e. SV or FMV) is used to reduce hyper-toned muscle spasticity and improve function in the paretic lower limbs of chronic stroke patients ^49^. In the upper limbs it, has been suggested that FMV acts to reduce spasticity in the stroke patient population ^50,51^. FMV studies have also reported improved gait performance in foot drop or calf muscle spasticity. FMV is suggested to be an effective complementary therapy for gait rehabilitation in stroke patients ^50,52^. However, FMV with different vibration stimulation frequencies tends to have different effects on flaccid and spastic muscle tones in patients of all three recovery stages (Acute-Chronic) of stroke ^49^. For example, rehabilitation programs that included a higher vibration frequency led to increased muscle strength and decreased spasticity and pain in people with chronic stroke ^53^. Thus, studies are needed to establish the minimum and optimal dose of FMV and its vibration stimulation characteristics, necessary to reach an excitatory response that could lead to a therapeutic benefit ^49^.

Overall, FMV is a modality of high interventional efficiency with significantly low cost. It is easier to administer and safe to use in stroke patients with no side effects and thus can be administered on the bedside of a sub-acute or acute stroke patient in rehabilitation or in homes for chronic stroke patients. However, majority of the evidence of FMV’s effects is evaluated in people with chronic or acute stroke ^54^. Clinical benefits of the sub-acute phase are yet to be determined.

The structural and functional changes that occur in the sub-acute stroke phase are crucial for the sensorimotor recovery in stroke patients ^22,55^. In addition, there is no consensus on the minimum dosage and optimal frequency of FMV stimulation to treat people with stroke. Further, many of the studies investigating the effects of FMV in stroke rehabilitation, use FMV device which is not wearable but is fixed at bedside. This may limit its access and effectiveness. The effects and effectiveness of wearable FMV when the FMV intervention is delivered over longer duration (> 4 weeks) are still unknown. In addition, only a handful of studies have investigated the effects of FMV on the upper limb function. The majority of studies have focused on the effects of FMV on limb spasticity rather than the overall function of the extremity ^51,56^.

Therefore, this study aimed to assess:

1. whether eight weeks of wearable FMV combined with conventional physical therapy had a greater impact on upper limb impairment and function than conventional physical therapy alone, in people with subacute stroke,
2. the overall recovery of upper limb motor functions rather than only spasticity as an outcome. This was assessed via FMUE and a Motor assessment test, in people with subacute stroke,
3. the effects of two FMV stimulation frequencies (60 Hz, 120 Hz), combined with conventional physical therapy, on upper limb impairment and function in people with sub-acute stroke.

## 2 Materials and Methods

### 2.1 Study design and setting

The study was a parallel-group, randomized controlled trial (RCT) conducted at the Rehabilitation Centre of Railway General Hospital, Rawalpindi, Pakistan, from February 26, 2020, to April 20, 2021. The Ethical Review Committee of Riphah International University, Pakistan, approved the study (REC/00654). All procedures performed in studies involving human participants followed the ethical standards of the institutional research committee and adhered to the declaration of Helsinki. The study was registered on the National Institutes of Health ClinicalTrials.gov clinical trial registry (NCT04289766). An information sheet was given to participants, and all participants signed a written consent form in presence of the researcher before the experiment.

### 2.2 Study participants

Participants were recruited from the rehabilitation department database of Railway General Hospital, Pakistan by contacting the potential participants via telephone and inviting them to participate. Participants were included if they were aged between 35 and 65 years, had sustained stroke more than six weeks and less than 12 weeks ago, had less than 53 on a upper extremity Fugl-Meyer Assessment (FMUE) of motor function (i.e., they had significant motor impairment)^57–59^, and the spasticity score for Bicep Brachialis and Extensor Carpi Radialis muscle of affected side was <3 on Modified Ashworth scale (MASh). Participants were excluded if they had any other neurological deficit, diabetic ulcer, infection or amputation of the limb, serious cardiovascular disease or unstable angina, serious orthopaedic problem, or chronic medical problems.

### 2.3 Intervention

During the eight-week intervention period, conventional physical therapy was delivered three days per week (a total of 24 sessions). The two experimental groups received the conventional physical therapy (the same as control group) for the same amount of time, in addition to FMV. The FMV was delivered after the conventional physical therapy for 10 minutes to both targeted muscles simultaneously, Biceps Brachii (BB) and Extensor Carpi Radialis (ECR) of the paretic limb. Each session took place under the supervision of experienced and certified physical therapist.

#### 2.3.1 Conventional Physical Therapy (PT Group, aka Group-1)

The conventional physical therapy group (group 1), here onwards simply referred as physical therapy group, underwent three comprehensive sessions of physical therapy per week for eight weeks, with an estimated duration of 40 minutes each. The physical therapy program consisted of muscle stretching and strengthening, balance exercises in sitting and standing positions, sit to stand practice, transfer practice according to patient needs, walking, stair climbing, upper limb functional training (reach, grasp, and hand to mouth activities), muscle tone inhibition techniques, postural stability control, sensory techniques, and daily functional activities. Hot packs and TENS were used to reduce pain or for muscle relaxation if required ^57,59^. Also, the participants were encouraged to continue performing exercises at home. The physical therapist that delivered the physical therapy treatment had five years of experience treating patients with neurological disorders.

#### 2.3.2 60 Hz Wearable Focal Muscle Vibration (PT + FMV60 Hz, aka Group/Group 2)

Wearable Focal Muscle Vibration (see 2.3.4 for more details) was applied to the paretic limb muscles with increased tone (i.e., Bicep Brachialis and Extensor Carpi Radialis) at a frequency of 60 Hz in addition to physical therapy.

#### 2.3.3 120 Hz Wearable Focal Muscle Vibration (PT + FMV120 Hz, aka Group/Group-3)

Wearable Focal Muscle Vibration (see 2.3.4 for more details) was applied to the paretic limb muscles with increased tone (i.e., Bicep Brachialis and Extensor Carpi Radialis) at a frequency of 120 Hz in addition to physical therapy.

#### 2.3.4 Wearable Focal Muscle Vibration Device and Delivery Details

For both Group 1 and 2 above, custom developed wearable vibration device capable of generating precise vibration frequencies of 60 Hz and 120 Hz with a constant vibration amplitude of 0.5mm (peak to peak) was used to deliver the FMV. The FMV device was worn (through a wearable sleeve) on the muscle belly of the target muscle, near its distal tendon insertion, so that sinusoidal vibration stimuli were delivered perpendicular to the muscle body. FMV was administered by a single experienced physiotherapist to minimize any variation in the location and tautness of the vibration therapy application ensuring good mechanical contact of the vibration actuator with the muscle.

### 2.4 Outcome Measures

Primary outcome measures included Fugl Meyer Scale for Upper Extremity (FMUE), Motor Assessment Scale (MAS), and Modified Ashworth Scale (MASh).

#### 2.4.1 Fugl Meyer Scale for Upper Extremity (FMUE)

Motor domain of the FMUE was used to assess post-stroke upper limb motor function. FMUE is subscale of Fugl-Meyer Assessment scale which is a valid and reliable performance-based impairment scale recommended to quantify recovery after stroke ^57,58^. FMUE consists of 33 items that include assessment of movement ability, coordination, speed, control and reflex activity ^60^. The scoring ranges from 0 to 66 such that higher FMUE score indicates less impairment of the affected upper limb ^59^.

#### 2.4.2 Motor Assessment Scale (MAS)

The MAS is a performance-based scale that evaluates everyday motor function in people with stroke. It consists of eight items that assess eight motor function tasks. These include supine to side lying, supine to sitting over the edge of a bed, balanced sitting, sitting to standing, walking, upper arm function, hand movements and advanced hand activities. Each task was performed three times and the best performance was used for analysis. The eight items are assessed using a 7-point scale (0 to 6). In addition, general tonus was assessed to provide an estimate of muscle tone on the affected side. The scoring ranges from 0 to 54. A higher score indicates better upper limb function^59^.

#### 2.4.3 Modified Ashworth Scale (MASh)

The MASh is the most widely used tool to assess muscle tone and has greater reliability for the upper limb. It is 5-point scale where participants are scored from 0 (no increase in muscle tone) to 4 (affected part rigid in flexion or extension) ^61^.

All outcome measures were assessed at the baseline and after eight weeks of intervention.

### 2.5 Randomization and Blinding

Randomization was carried out following the baseline assessment using the sealed envelope method. All participants and the outcomes assessors were blinded to group allocation. The statistician who analysed the data was also blinded to group allocation, as all recorded data were anonymized and coded before being provided for analysis. The physical therapists providing the physical therapy intervention could not be blinded to group allocation.

### 2.6 Statistical Analysis

The null hypothesis for the statistical analysis was that there is no difference in any of the outcome measures (FMUE, MAS, and MASh) across the intervention groups at the post-intervention time-point. A separate linear regression model for longitudinal analysis of covariance was set up for each outcome to evaluate this hypothesis. For FMUE and MAS, the model regresses the post-experiment outcome scores on the intervention groups (discrete; Control (PT), PT + FMV60 Hz and PT+ FMV120 Hz) while adjusting for baseline outcome scores and pre-specified co-variates including age (continuous), time since stroke (continuous), sex (dichotomous) and the affected side (dichotomous). For MASh, the model regresses rank changes on the same explanatory variables. The rank changes were obtained by subtracting post-experiment scores from pre-experiment scores. Therefore, MASh scores of 3 at baseline and 0 at post-experiment imply a rank change of positive 3. A sensitivity analysis for the robustness of this mean rank change linear model was also conducted by fitting a proportional odds model (a.k.a. ordinal regression). This model estimates the odds ratios for higher rank change corresponding to the experiment groups and the pre-specified co-variate variables. Only the mean rank change linear model results are reported if both models lead to the same results. The goodness-of-fit for the linear regression models was evaluated by inspecting the normality and homogeneity of variance of its residuals using QQ-plot, fitted-values versus residuals plot and histogram. The null hypothesis was evaluated with an analysis of variance test conducted on the fitted linear regression models, which compares the outcome-variance explained by variable representing groups to error-variance to determine statistical significance. The mean estimates for the outcomes across groups are reported along with their 95% confidence intervals. Pair-wise mean differences (contrasts) are also reported along with their 95% confidence intervals and tests for the null hypotheses that each pair difference is zero if the main effect is significant. The statistical significance level was set at 0.05. The statistical analysis was conducted in R (The R Foundation for Statistical Computing, 4.1.0) using packages car (version 3.0-10), MASS (version 7.3-54) and ggplot2 (version 3.3.3) ^62–65^.

## 3 Results

Of the 138 volunteers who were assessed for eligibility, 98 met the inclusion criteria and agreed to participate in the study. Ninety participants completed the 8-week intervention and were assessed post-intervention, n= 30 in the Group 1 (PT group), n= 30 in the Group 2 (PT+FMV60 Hz), n= 30 in the Group 3 (PT+FMV120 Hz). Although, participants were allocated to these three groups randomly, the group numbers (n = 30) were balanced to achieve viable comparison between them. Eight participants dropped out of the study due to transportation limitations to the hospital where the study was being conducted. The study flow is show in Figure 1.

**Figure 1:**
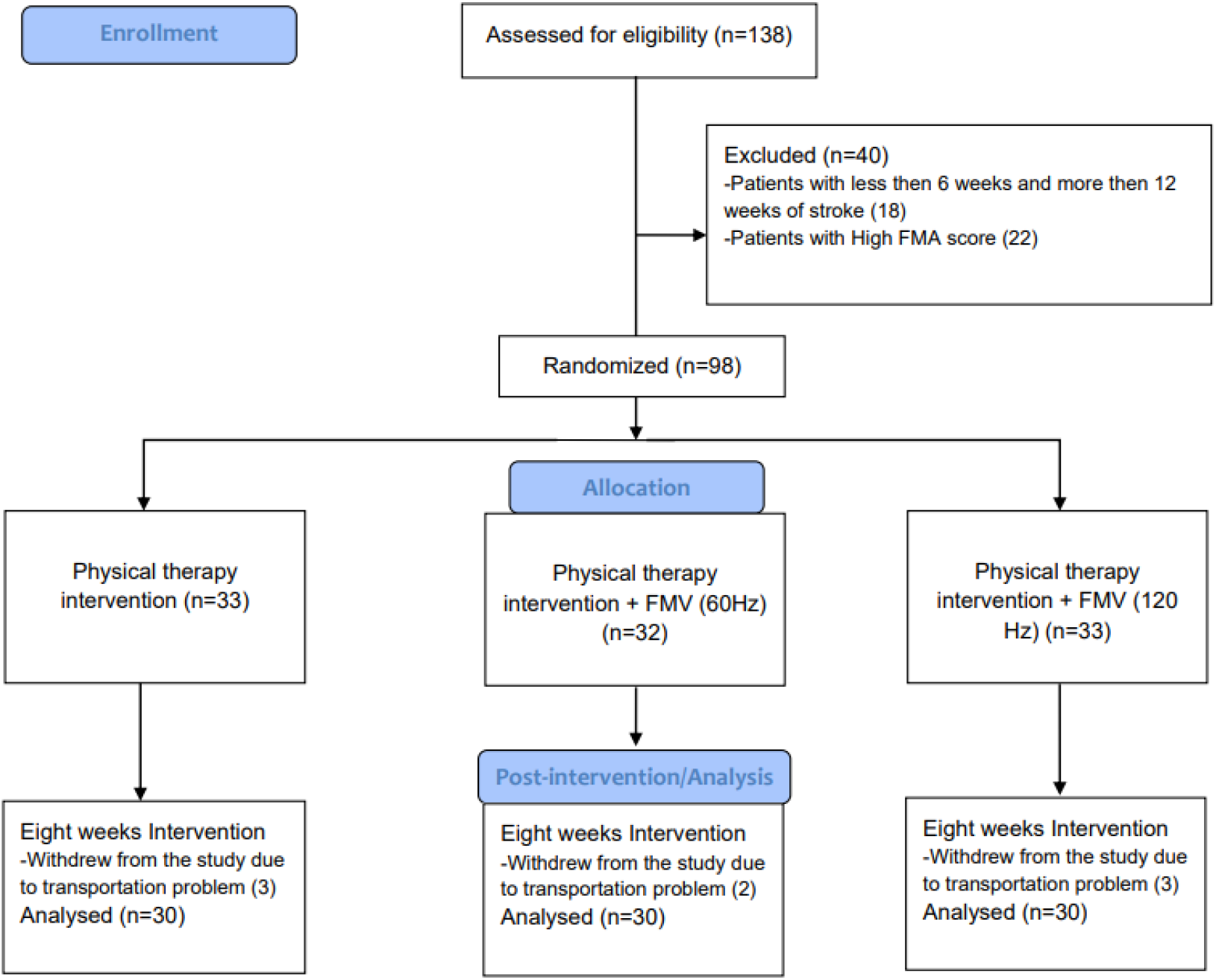
Flow and classification of interventions.

The clinical characteristics of participants are given in Table 1.

**Table 1.** Clinical characteristics of participants in each group.

| **Variables** | **PT** | **PT + FMV (60 Hz)** | **PT + FMV (120 Hz)** |
| --- | --- | --- | --- |
| **Gender** |  |  |  |
| Male (n) | 18 | 20 | 20 |
| Female (n) | 12 | 10 | 10 |
| **Age, years (mean +/- SD)** | 51 +/- 6.67 | 50.53 +/- 11.13 | 50.56 +/- 10.36 |
| **Side of body affected by stroke** |  |  |  |
| Left (n) | 13 | 15 | 15 |
| Right (n) | 17 | 15 | 15 |

n: number of participants, SD: standard deviation, PT: Physical Therapy, FMV: Focal Muscle Vibration.

### 3.1 Between-group differences

At post-intervention time-point, there was statistically significant difference in FMUE scores between the three groups (F (2, 81) = 7.2, p = 0.001). Compared to the PT group, both PT + FMV60 Hz and PT + FMV120 Hz groups had significantly higher FMUE mean scores (p < 0.05). However, there were no significant differences between PT + FMV60 Hz and PT + FMV120 Hz groups (p = 0.5). The PT + FMV120 Hz group had the highest post-intervention mean change score (49.9 95% CI [48.5, 51.3]).

Although there was no statistically significant difference between PT + FMV60 Hz and PT + FMV120 Hz, the pre-to post-intervention changes above overall mean change (FMUE = 17.3, MAS = 8.4, MASh = 1.3), the number of stroke people crossing this threshold are more in the group treated with PT + FMV120 Hz as compared to PT + FMV60 Hz (Figure 2).

**Figure 2:**
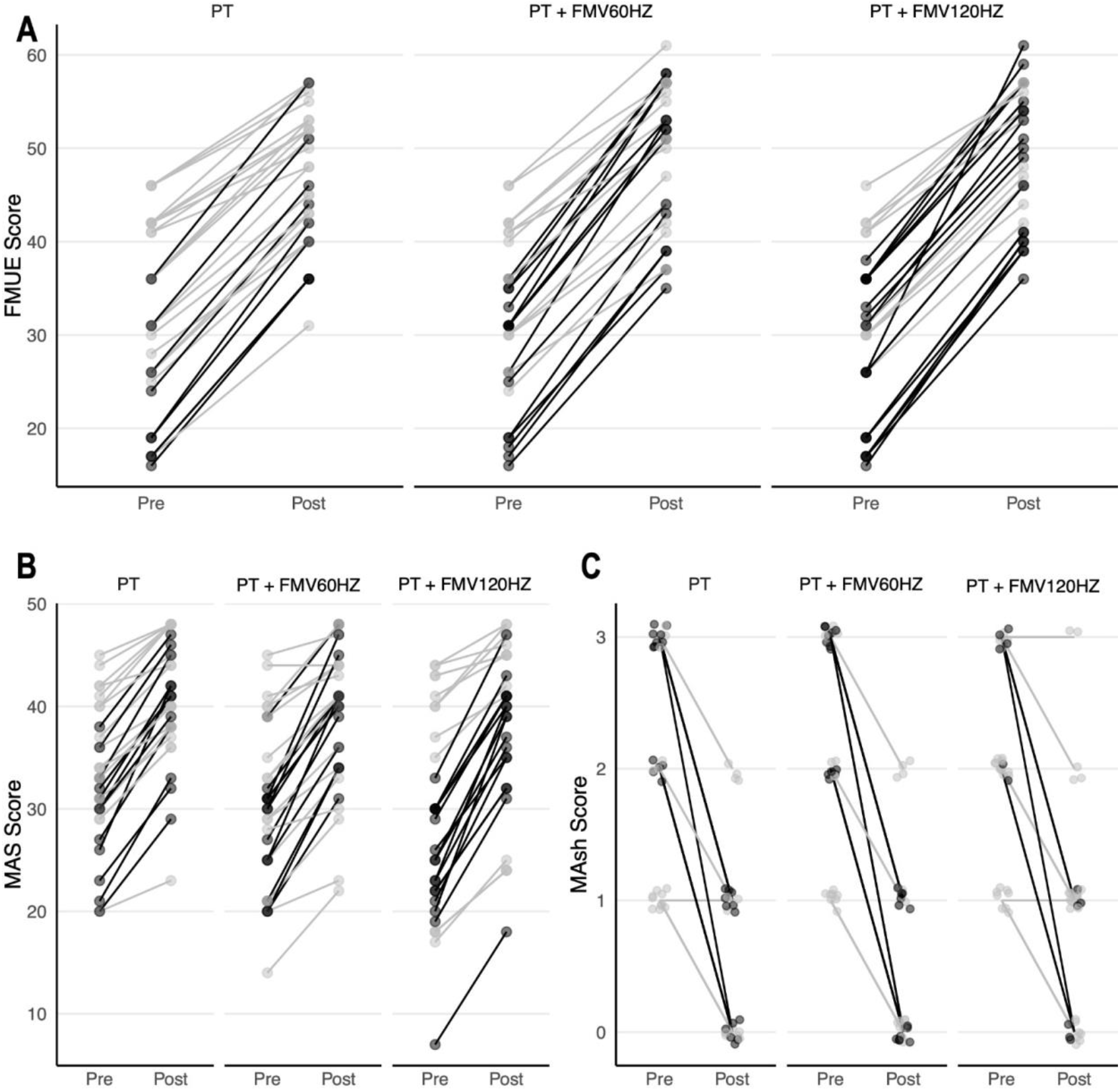
Participant-wise FMUE and MAS scores, for the three interventions at pre-randomization baseline and the end of the interventions (8 weeks). These are based on pre-to post-difference thresholds: MAS: 8.36, MASh: 1.35. and FMUE:17.28. Any change greater than the threshold is printed in dark shade whereas smaller changes are greyed out. Note: FMUE: Fugl-Meyer assessment of upper extremity, PT: physical therapy and FMV: Focal Muscle Vibration.

There was no statistically significant difference in MAS scores between the three groups (F (2, 81) = 0.2, p = 0.8). All groups had similar post-intervention mean change scores.

There was statistically significant difference in MASh rank changes between the three groups (F (2, 81) = 3.3, p = 0.04). PT + FMV60 Hz group had the highest mean rank change (1.5 95% CI [1.3, 1.8]), and PT + FMV120 Hz group had the lowest mean rank change (1.2 95% CI [1.0, 1.4]) at the post-intervention time-point. The difference in mean rank change between these two groups was statistically significant (p=0.02)). However, there were no statistically significant differences when the PT group was compared with PT + FMV60 Hz group or PT + FMV120 Hz group.

The mean estimates for between-group differences and within-group differences are listed in Table 2 and 3, respectively.

**Table 2.** Between-group differences in means and mean rank changes at the post-intervention time-point.

| Outcome | Group Contrast | Difference ± S.E. [95% CI] | H<sub>0</sub>: Difference = 0<br>T-value [df = 81], P-Value |
| --- | --- | --- | --- |
| FMUE | 1 – 2 | -2.7 ± 0.9 [-4.6, -0.9] | -2.9, 0.005 * |
| | 1 – 3 | -3.4 ± 0.9 [-5.3, -1.5] | -3.6, 0.0006 * |
| | 2 – 3 | -0.6 ± 0.9 [-2.5, 1.3] | -0.7, 0.5 |
| MAS | 1 – 2 | 0.0 ± 0.9 [-1.8, 1.8] | |
| | 1 – 3 | -0.5 ± 0.9 [-2.3, 1.4] | |
| | 2 – 3 | -0.5 ± 0.9 [-2.3, 1.3] | |
| MASh | 1 – 2 | -0.1 ± 0.1 [-0.4, 0.2] | -0.7, 0.5 |
| | 1 – 3 | 0.3 ± 0.1 [0.0, 0.6] | 1.8, 0.08 |
| | 2 – 3 | 0.4 ± 0.1 [0.1, 0.7] | 2.5, 0.02 * |

**Note:** “\*” denotes statistical significance ( $p < 0.05$ ). SE stands for standard error, and CI stands for the confidence interval. Group 1: PT, Group 2: PT + FMV60 Hz, Group 3: PT+ FMV120 Hz.

**Table 3.** Group means and mean rank changes at the post-intervention time-point estimated by linear regression model by adjusting for pre-intervention scores, age, sex, affected-side and time-since-stroke.

| Outcome | Group | Mean ± S.E. [95% CI] |
| --- | --- | --- |
| FMUE | PT | 46.5 ± 0.7 [45.2, 47.9] |
| | PT + FMV60 Hz | 49.3 ± 0.7 [47.9, 50.6] |
| | PT + FMV120 Hz | 49.9 ± 0.7 [48.5, 51.3] |
| MAS | PT | 38.9 ± 0.6 [37.6, 40.1] |
| | PT + FMV60 Hz | 38.9 ± 0.7 [37.6, 40.2] |
| | PT + FMV120 Hz | 39.3 ± 0.7 [38.0, 40.7] |
| | | Mean Rank Change ± S.E. [95% CI] |
| MASh | PT | 1.4 ± 0.1 [1.2, 1.6] |
| | PT + FMV60 Hz | 1.5 ± 0.1 [1.3, 1.8] |
| | PT + FMV120 Hz | 1.2 ± 0.1 [1.0, 1.4] |

**Note:** “\*” denotes statistical significance ( $p < 0.05$ ). SE stands for standard error and CI stands for the confidence interval. PT: Physical Therapy, FMV: Focal Muscle Vibration.

The outcome data from participants is shown in **Error! Reference source not found**..

## 4 Discussion

This is the first study to evaluate the combined effects of FMV and conventional physical therapy on the upper limb motor function in people with sub-acute stroke. It is also the first study of FMV on sub-acute stroke cohort of larger size (∼n = 90), especially where the effects of FMV, delivered through wearable FMV device, are tested over 8 weeks duration. It is also the first study to compare the effects of two FMV vibration stimulation (60 Hz and 120 Hz) frequencies over the period of 8 weeks duration in stroke population.

It was found that wearable FMV combined with physical therapy significantly improved upper limb motor function (as judged through FMUE) compared to physical therapy alone, irrespective of the FMV stimulation frequency. However, there was no significant difference in everyday motor function (as judged through MAS) between the three groups. In addition, PT + FMV60 Hz group showed significant reduction in upper limb spasticity (as judged through MASh) as compared to PT + FMV120 Hz group, while there was no significant change in spasticity when the PT group was compared with PT + FMV60 Hz group or PT + FMV120 Hz group.

### Effects on the Upper Limb Function

Eight weeks of PT+FMV, irrespective of FMV frequency, improved upper limb motor function in comparison to PT (alone) group. Previous studies, comparing a program of PT+FMV with PT alone, have shown that two weeks of PT + FMV at 120 Hz improved paretic upper limb kinematics ^23^ and modulated upper limb muscle activity during the reaching movements ^66^ in people with chronic stroke. Improvements in lower limb gait function have also been noted following four weeks of PT+FMV120 Hz in people with chronic stroke ^49^. So far only one single group, pre-post study has evaluated the effects of single session of FMV at 60 Hz and found improved manual dexterity in people with chronic stroke with spastic hemiparesis ^67^. Thus, previous studies support our results that the use of FMV in addition to PT improve functional capacity in stroke patients. However, our study shows for the first time that PT+ FMV intervention can be beneficial in improving upper limb function in sub-acute stroke stage too. It also shows that delivering FMV through a wearable FMV device, over longer duration is feasible and effective in improving upper limb function in stroke.

### Effects on Spasticity

Although eight weeks of PT+FMV (both 60 HZ or 120 Hz) did not significantly change upper limb spasticity in comparison to PT alone, PT+FMV at 120 Hz significantly reduced upper limb spasticity more than PT+FMV at 60 Hz. In contrast, previous studies in people with chronic stroke have found reduction in upper limb spasticity following eight weeks of PT+FMV30Hz ^50^ and two weeks of PT+FMV at 100 Hz ^68^ and 120 Hz ^66^ when compared to PT alone or placebo + PT. Other post-stroke studies using single session of FMV have also found reduction in spasticity when delivered at 91 Hz ^56^ and 300 Hz ^69^. The differences in spasticity reduction between studies in chronic stroke people compared to our study in sub-acute stroke people can be attributed, at least in part, to the ongoing spontaneous motor recovery from acute to chronic phase of the stroke. According to Brunnstrom ^70^ evolution of motor recovery and emergence and eventual disappearance of spasticity occurs parallelly throughout various phases of the stroke (acute, sub-acute, chronic) ^43,70^. Thus, whether no change in spasticity observed in this study was due the sub-acute nature of stroke where spasticity is still developing or whether the improvements in spasticity caused by FMV were not statistically significant enough is unclear.

Although there was no statistical significant difference between two frequencies (60 and 120 Hz), however if we see pre-to post-intervention changes above overall mean change (FMUE = 17), there is a trend that more patient cross this threshold in group treated with PT + FMV120 Hz as compared to PT + FMV60 Hz. Frequency of FMV stimulation is crucial, as the is evidence suggests that Ia-afferents can fire synchronously with vibration frequencies up to 80–120 Hz ^55,71^. Differences in the results from both the frequencies give a better perspective of frequencies to be used for rehabilitation.

### Potential Underlying Neural Mechanisms

FMV produces strong proprioceptive stimulus reaching not only spinal but supraspinal structures. At peripheral-spinal level, it is well known that FMV entrains muscle spindle afferent firing rate to stimulation frequencies between 80-120 Hz ^19,21,22^. Further, recent evidence suggests that FMV creates a strong proprioceptive stimulus, which via 1a afferents, induces long-term depression like plasticity in relevant spinal cord circuits, perhaps indicating ability to induce plasticity at Ia-montoneuron synapse level ^72^.

At cortical level, entrainment of neuronal firing to FMV stimulation frequency, augments proprioceptive input to the CNS. Studies in both healthy and stroke population have shown increase in motor evoked potentials (MEP) when FMV was delivered to the upper limb flexor muscle^21,22^. Further, FMV increased MEPs in the vibrated muscle and decreased MEPs in the non-vibrated muscle ^73,74^. When FMV was applied for the longer duration (∼60 minutes), increase in MEPs recorded from flexor muscle was seen for up to 60 minutes following FMV stimulus ^75,76^. Imaging studies have shown increases in FMV stimulation related activities in motor cortex, premotor cortex, and supplementary and cingulate motor areas ^77–79^. Thus, although the exact underlying mechanisms responsible for cortical projections are not clear it can be posited that strong proprioceptive stimulus generated by FMV through Ia afferents, has ability to increase cortical activation and corticomotor drive, which through a Hebbian like synaptic plasticity mechanism, may be responsible for improvement seen in functional outcomes, including seen this study.

Regarding spasticity, its development shortly follows stroke ^80^ while neuronal plastic changes usually taking place between 1-6 weeks ^81^. These plastic changes are usually manifested as muscle overactivity and hyper reflex resulting in spasticity ^43,82^. In a longitudinal study examining the development of spasticity over 6 weeks after stroke, found patients who recovered arm function showed signs of spasticity at all assessment points^83^. However, patients who did not recover useful arm function had signs of spasticity over the course of a 36-weeks follow-up. In this study we did not follow patients up, as such we were unable to establish which of the patients went on to develop lasting spasticity and did not recover their arm function and which ones recovered their arm function and had limited lasting spastic symptoms. However the above evidence underscores the complexity of investigating changes in spasticity during acute stroke phase. Due to ongoing spontaneous recovery in this phase, it is challenging to differentiate the effects of intervention such as FMV on spasticity.

Further, many FMV studies which have reported reduction in spasticity have delivered FMV stimulation to the antagonist-extensor muscle, based on the evidence that application of vibration stimulation to the antagonist significantly reduces hyper reflex activity in agonist-flexor muscle through reciprocal inhibition, in turn positively modulating flexor muscles activity and strength ^84^. Whereas, some studies have delivered FMV to the agonist-flexor muscle and still saw reduction in spasticity in chronic stroke patients ^22,85^. However, Marconi et al., also asked their stroke participants to maintain voluntary contraction of their paretic limb, to which agonist-flexor muscle, the FMV was applied. Marconi et al., reasoned that ongoing muscle contraction induced descending volley, when combined with ascending volleys (induced by FMV afferent stimulation), potentially induce long-lasting changes in corticomotor excitability of the target muscle (i.e. inhibition) as well as of its functional antagonist (i.e. facilitation). Thus, exact mechanisms responsible for reduction in spasticity upon application of FMV to either agonist or antagonist muscle, need further investigation. This, along with, the ongoing recovery during acute phase of the stroke, might explain why we did not see reduction in spasticity following FMV intervention.

### 4.1 Limitations

The main limitation of this study is the lack of a follow-up assessment, thus depriving us of long-term recovery results. Further, the population may be considered relatively young for stroke, and younger patients are likely to have a better recovery. However, in Pakistan where the study was carried out, 20% of strokes ^86^ occur in patients less than 45 years of age, where the average life span is close to 70 years.

Results of this study show that FMV has a prominent effect on the motor recovery of the upper limb, as shown by the FMUE outcome scores. FMVs of constant and 60-120 Hz are considered safe, easy, and well-tolerated intervention. Thus, it can be used in addition to conventional physiotherapy as an intervention to promote upper limb function in sub-acute stroke patients.

## 5 Conclusion

a. This study provided the first evidence that delivering long-term (> eight weeks) FMV through wearable FMV device is not only feasible but also an effective intervention to improve upper limb function, from sub-acute stroke phase onwards.
b. Compared to the control group, both 60 and 120 Hz of wearable FMV improved upper limb motor function when these interventions were added to eight weeks of conventional physical therapy in people with sub-acute stroke.
c. Although not statistically significant, more stroke people with 120 Hz FMV intervention showed greater changes compared to the 60 Hz FMV intervention. Perhaps signifying 120 Hz to be more effective FMV stimulation frequency of the two, in improving upper limb function.
d. Compared to the control group, both 60 and 120 Hz of wearable FMV did not significantly improve spasticity outcomes for people with sub-acute stroke. Whether smaller change in spasticity outcome was due the sub-acute stage of stroke where spasticity is still developing or whether the changes in spasticity were not statistically significant enough is unclear.

To summarize, wearable FMV has the potential to augment the rehabilitative benefits of convention physical therapy in people with sub-acute stroke. Further research, involving larger FMV dose (frequency of intervention and time) and follow-up assessment(s), is required to build on these findings. Longer -term impacts (> 6 months) of FMV on motor function in people with stroke also needs investigation to substantiate clinical relevance of these finding.

## Data Availability

Data will be available upon request.

## 6 Data Availability

Data will be available upon request.

## 7 Acknowledgments

We would like to thank the participants involved in the study for their time and effort.

## 8 Funding

Partial funding support for this study was though the Royal Academy of Engineering (RAEng)-Leverhulme Trust Fellowship awarded to ANP.

## 9 Competing Interests

None declared.

## Notes

### Competing Interest Statement

The authors have declared no competing interest.

### Clinical Trial

NCT04289766

